# Global research priorities for COVID-19 in maternal, reproductive and child health: results of an international survey

**DOI:** 10.1101/2021.07.13.21260207

**Authors:** Melanie Etti, Jackeline Alger, Sofía P. Salas, Robin Saggers, Tanusha Ramdin, Margit Endler, Kristina Gemzell-Danielsson, Tobias Alfvén, Yusuf Ahmed, Allison Callejas, Deborah Eskenazi, Asma Khalil, Kirsty Le Doare

## Abstract

**Background:** The World Health Organization’s “*Coordinated Global Research Roadmap: 2019 Novel Coronavirus*” outlined the need for research that focuses on the impact of COVID-19 on pregnant women and children. More than one year after the first reported case, significant knowledge gaps remain, highlighting the need for a coordinated approach. To address this need, the Maternal, Newborn and Child Health Working Group (MNCH WG) of the COVID-19 Clinical Research Coalition conducted an international survey to identify global research priorities for COVID-19 in maternal, reproductive and child health.

**Method:** This project was undertaken using a modified Delphi method. An electronic questionnaire was disseminated to clinicians and researchers in three different languages (English, French and Spanish) via MNCH WG affiliated networks. Respondents were asked to select the five most urgent research priorities among a list of 17 identified by the MNCH WG. Analysis of questionnaire data was undertaken to identify key similarities and differences among respondents according to questionnaire language, location and specialty. Following elimination of the seven lowest ranking priorities, the questionnaire was recirculated to the original pool of respondents. Thematic analysis of final questionnaire data was undertaken by the MNCH WG from which four priority research themes emerged.

**Results:** Questionnaire 1 was completed by 225 respondents from 29 countries. Questionnaire 2 was returned by 49 respondents. The four priority research themes were 1) access to healthcare during the COVID-19 pandemic, 2) the direct and 3) indirect effects of COVID-19 on pregnant and breastfeeding women and children and 4) the transmission of COVID-19 and protection from infection.

**Conclusion:** The results of these questionnaires indicated a high level of concordance among continents and specialties regarding priority research themes. This prioritized list of research uncertainties, developed to specifically highlight the most urgent clinical needs as perceived by healthcare professionals and researchers, could help funding organizations and researchers to answer the most pressing questions for clinicians and public health professionals during the pandemic. It is hoped that these identified priority research themes can help focus the discussion regarding the allocation of limited resources to enhance COVID-19 research in MNCH globally.

## Introduction

More than one year has passed since the first case of COVID-19 was reported on 31^st^ December 2019, leading to the declaration of a pandemic by the World Health Organization (WHO) on 11^th^ March 2020. Since its emergence, there has been a surge of research conducted to understand how COVID-19 affects pregnant women, their developing fetuses and children. The WHO report entitled “*A Coordinated Global Research Roadmap: 2019 Novel Coronavirus*” outlined the need for research that focuses on the impact of COVID-19 on pregnant women and children, however, there are still considerable gaps in this knowledge.(1) Much of the data relating to COVID-19 has been derived from research conducted in high-income countries (HICs), producing findings that may not be relevant or generalizable to women and children in low-and middle-income countries (LMICs). Furthermore, health systems and public health strategies used to control the spread of the virus in different parts of the world may vary according to the availability of funding and resources, meaning a “one-size-fits-all” approach cannot be appropriately applied to determine research priorities in different global regions.(2,3)

In light of these varying needs, it is imperative that we strive to understand priorities in different international regions. Such an approach is necessary to target funding, resources and research efforts where needed, and to guide policy makers to ensure that efforts are made to address the most pressing research questions for pregnant women and children. To address these needs, the Maternal, Newborn and Child Health Working Group (MNCH WG) of the COVID-19 Clinical Research Coalition, which is comprised of obstetricians, gynecologists, public health physicians and pediatricians, conducted an international survey to identify the most pressing COVID-19 research priorities in maternal, reproductive and child health.(4) In pursuing this line of work, we sought to identify priorities that were equitable, internationally representative, and also representative of the diverse specialties caring for pregnant women and children globally. We also sought to complement other COVID-19 research priority setting exercises currently being undertaken in this field,(5,6) in order to provide a broader insight into the needs of women and children around the world.

## Methods

### Development and dissemination of questionnaires

This project was undertaken between October 2020 and January 2021 using a modified Delphi method (Figure 1). A short online questionnaire was designed by the MNCH WG through collaborative discussions guided by the WHO’s “*Coordinated Global Research Roadmap for the Novel Coronavirus*”.(1) Questionnaire respondents were asked to provide demographic information including specialty and location, and to select the five most urgent research priorities among a list of 17 which had been generated by the MNCH WG through literature review and expert opinion (Appendix 1). Space was also provided within the questionnaire for respondents to include additional research priorities that were not included in the list. The questionnaire was translated from English into French and Spanish and disseminated electronically using the SurveyMonkey platform via affiliated networks of the MNCH WG members including: World Society for Pediatric Infectious Diseases (WSPID); European Society for Pediatric Infectious Diseases (ESPID); African Society for Pediatric Infectious Diseases (AfSPID), who forwarded to their respective in-country pediatric and obstetric associations; Honduran Pediatric Association; Royal College of Obstetricians and Gynaecologists (RCOG) and Royal College of Paediatrics and Child Health (RCPCH), who forwarded to their global health membership; American College of Obstetricians and Gynecologists (ACOG) and International Federation of Gynecology and Obstetrics (FIGO), who also forwarded to all its country member organizations for further dissemination.(7–10) We also disseminated the Questionnaire 1 via the social media platforms Twitter and Facebook. Responses were collected electronically over a period of four weeks and data analysis was performed using Microsoft Excel.

**Figure 1.**
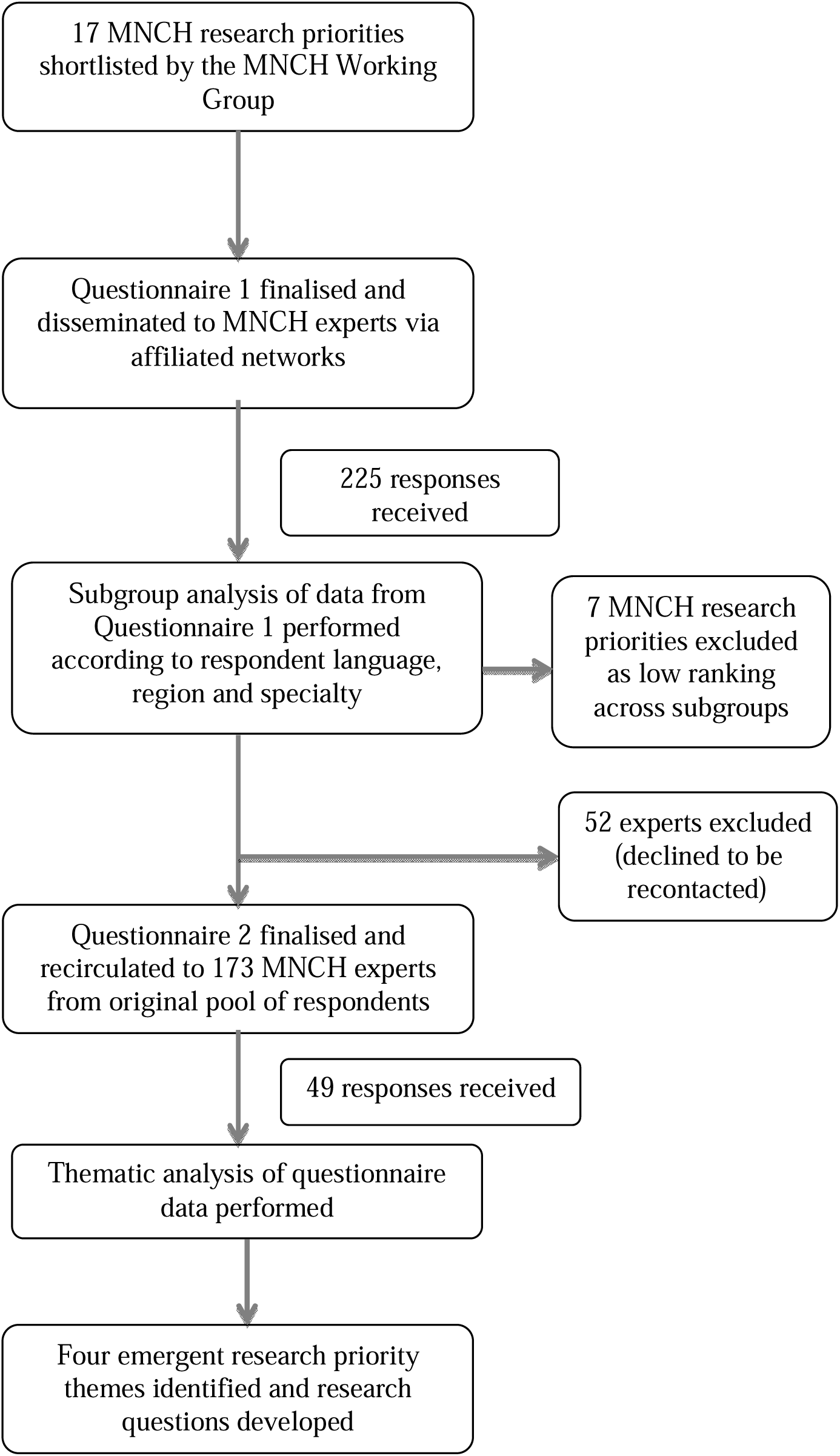
Overview of the modified Delphi method implemented for the development of the maternal, reproductive and child health COVID-19 research priority themes, October 2020–January 2021

### Data analysis

The frequency of selection of each research priority was calculated (with each research priority being equally weighted). Subgroup analysis of Questionnaire 1 data was performed to determine the frequency of selection of each research priority according to questionnaire language, respondent specialty and respondent location (continent). An overview of additional research priorities submitted by respondents was also undertaken by the MNCH WG to determine whether they were sufficiently distinct from the original listed priorities (no new priorities were added). Following this review, the seven least frequently selected research priorities by questionnaire respondents among the original list of 17 (as indicated by the seven lowest scores) were eliminated from the list, leaving ten research priorities which were reviewed and refined from the MNCH WG and recirculated to respondents to Questionnaire 1 who had consented to being recontacted (Appendix 2). Respondents were asked to rank these ten research priorities from most to least important (with 1 signifying the most important priority and 10 signifying the least important priority) and to comment on the relevance and comprehensibility of the research priorities. Responses to Questionnaire 2 were also collected over a period of four weeks. The mean ranking of each research priority was calculated by dividing the sum of the rankings for each research priority by the total number of respondents to Questionnaire 2. We subsequently performed a thematic analysis of the most frequently selected priorities from Questionnaire 1 and the highest ranked priorities among Questionnaire 2, through which four principal research priority themes emerged. The MNCH WG reconvened to define important research questions within each theme. Three key questions within each research priority theme were developed following literature review to identify persisting knowledge gaps. We then presented the research questions to OBGYN and pediatric specialists at virtual international meetings to ensure agreement among specialists within the field.

## Ethical consideration

This project was reviewed by the Public Health England Research Ethics and Governance Group. Given the consultative nature of the surveys, the project was deemed not to require full ethical review and was granted an exemption from the ethical approval process (https://www.gov.uk/government/organisations/public-health-england/about/research).

## Results

Questionnaire 1 was completed by 225 respondents from 29 countries across five continents (Figure 2). The full list of countries can be found in Appendix 3. The demographic characteristics of Questionnaire 1 respondents and most frequently selected research priorities according to questionnaire language, respondent specialty and location are shown in Table 1.

**Figure 2.**
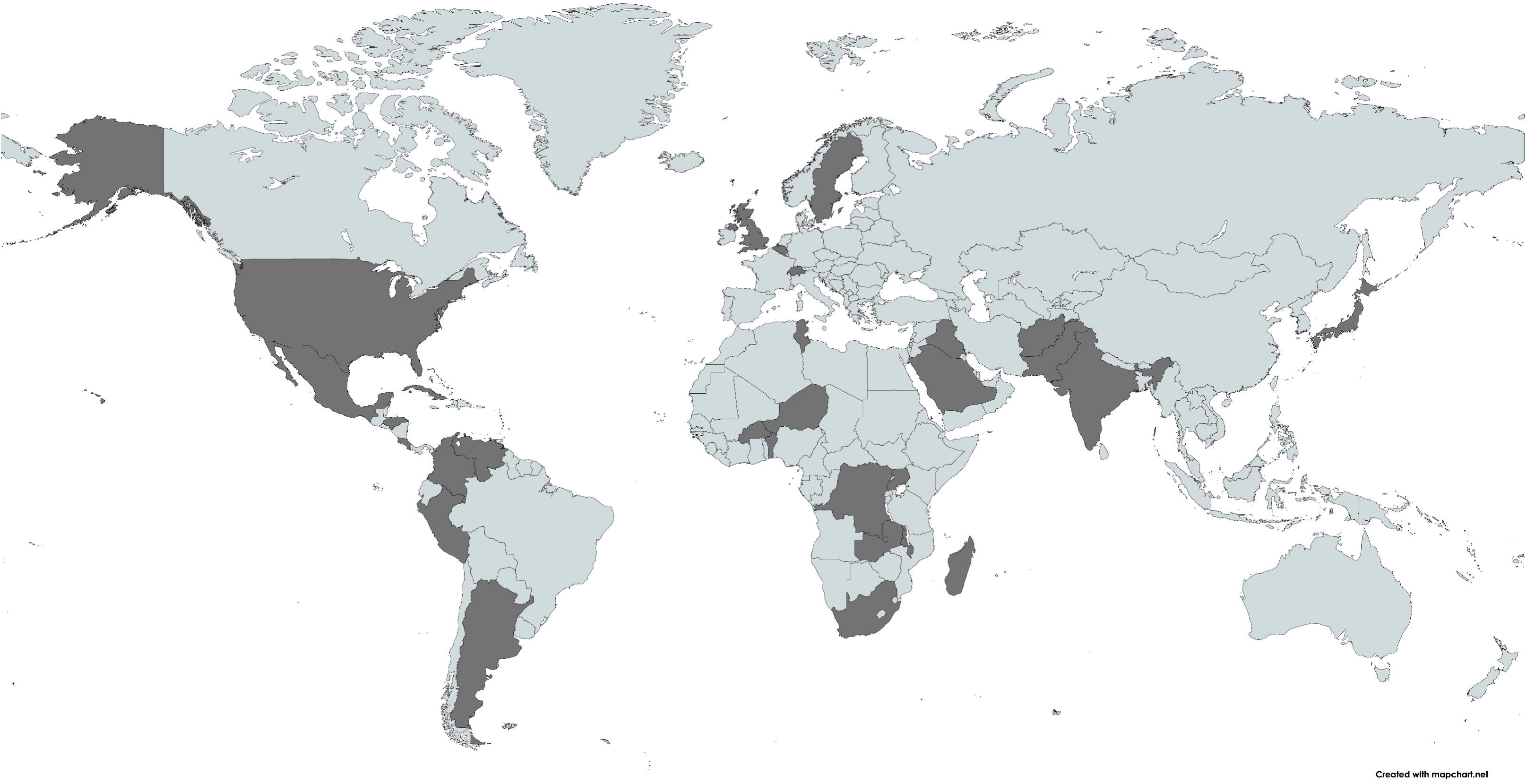
Country locations of Questionnaire 1 respondents

**Table 1.**
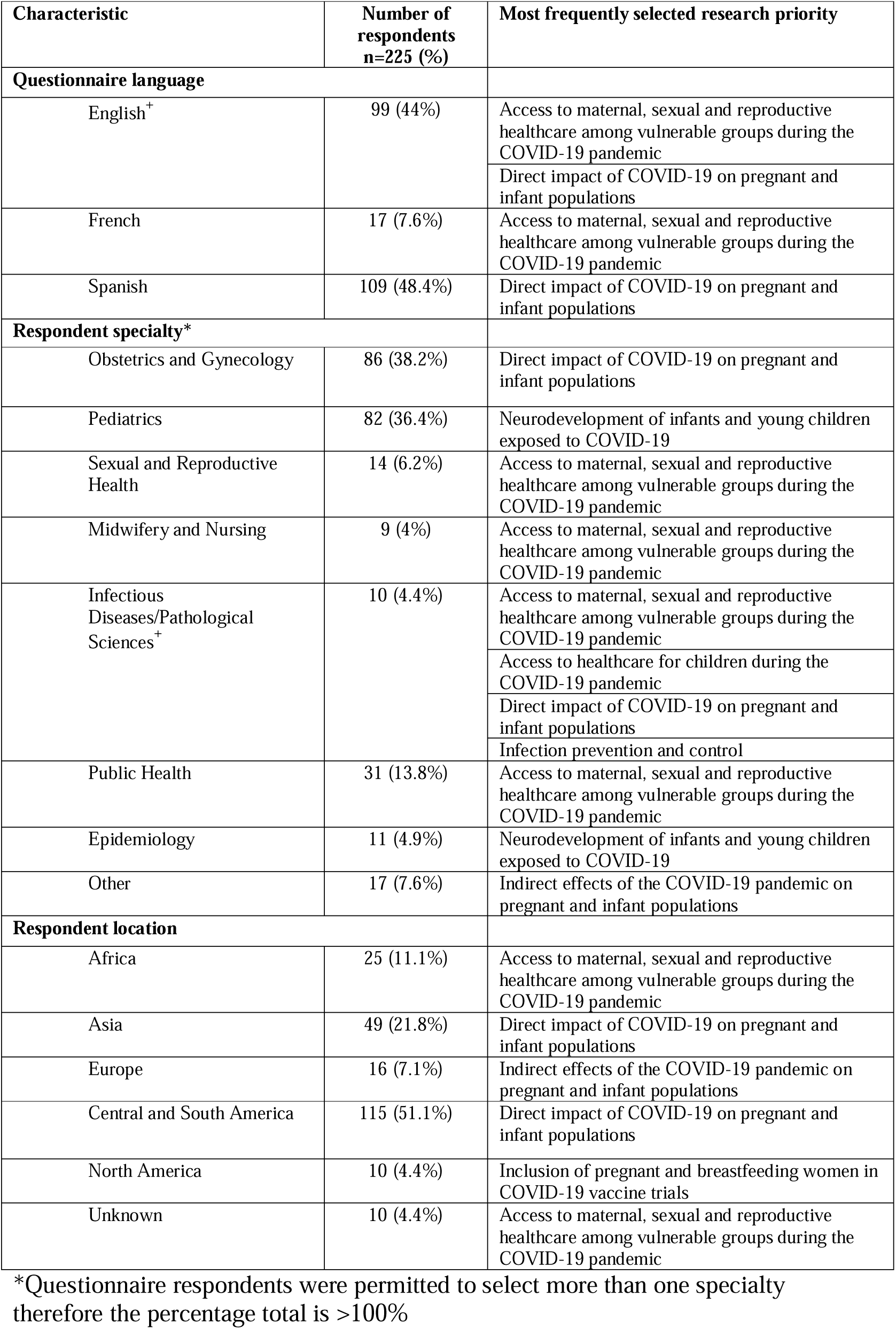
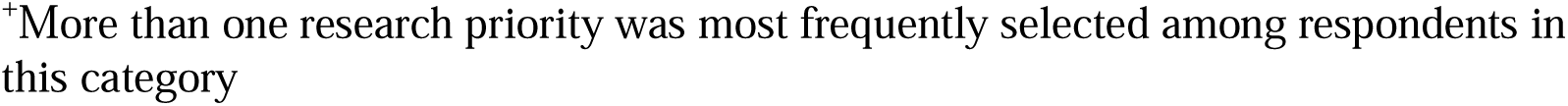
Demographic characteristics of Questionnaire 1 respondents and the most frequently selected research priorities according to questionnaire language, respondent specialty and location

| Characteristic | Number of respondents<br>n=225 (%) | Most frequently selected research priority |
| --- | --- | --- |
| **Questionnaire language** |  |  |
| English<sup>+</sup> | 99 (44%) | Access to maternal, sexual and reproductive healthcare among vulnerable groups during the COVID-19 pandemic |
|  |  | Direct impact of COVID-19 on pregnant and infant populations |
| French | 17 (7.6%) | Access to maternal, sexual and reproductive healthcare among vulnerable groups during the COVID-19 pandemic |
| Spanish | 109 (48.4%) | Direct impact of COVID-19 on pregnant and infant populations |
| **Respondent specialty\*** |  |  |
| Obstetrics and Gynecology | 86 (38.2%) | Direct impact of COVID-19 on pregnant and infant populations |
| Pediatrics | 82 (36.4%) | Neurodevelopment of infants and young children exposed to COVID-19 |
| Sexual and Reproductive Health | 14 (6.2%) | Access to maternal, sexual and reproductive healthcare among vulnerable groups during the COVID-19 pandemic |
| Midwifery and Nursing | 9 (4%) | Access to maternal, sexual and reproductive healthcare among vulnerable groups during the COVID-19 pandemic |
| Infectious Diseases/Pathological Sciences<sup>+</sup> | 10 (4.4%) | Access to maternal, sexual and reproductive healthcare among vulnerable groups during the COVID-19 pandemic |
|  |  | Access to healthcare for children during the COVID-19 pandemic |
|  |  | Direct impact of COVID-19 on pregnant and infant populations |
|  |  | Infection prevention and control |
| Public Health | 31 (13.8%) | Access to maternal, sexual and reproductive healthcare among vulnerable groups during the COVID-19 pandemic |
| Epidemiology | 11 (4.9%) | Neurodevelopment of infants and young children exposed to COVID-19 |
| Other | 17 (7.6%) | Indirect effects of the COVID-19 pandemic on pregnant and infant populations |
| **Respondent location** |  |  |
| Africa | 25 (11.1%) | Access to maternal, sexual and reproductive healthcare among vulnerable groups during the COVID-19 pandemic |
| Asia | 49 (21.8%) | Direct impact of COVID-19 on pregnant and infant populations |
| Europe | 16 (7.1%) | Indirect effects of the COVID-19 pandemic on pregnant and infant populations |
| Central and South America | 115 (51.1%) | Direct impact of COVID-19 on pregnant and infant populations |
| North America | 10 (4.4%) | Inclusion of pregnant and breastfeeding women in COVID-19 vaccine trials |
| Unknown | 10 (4.4%) | Access to maternal, sexual and reproductive healthcare among vulnerable groups during the COVID-19 pandemic |

\*Questionnaire respondents were permitted to select more than one specialty therefore the percentage total is >100%
<sup>†</sup>More than one research priority was most frequently selected among respondents in this category

Questionnaire 2 was disseminated to 173 members of our original pool of experts (52 declined to be recontacted) and was returned by 49 respondents. The research priority from Questionnaire 2 with the highest mean ranking was “Access to healthcare for children during the COVID-19 pandemic” which had an average score of 3.86/10. The lowest ranked priority was “Mental health sequelae of COVID-19 pandemic in pregnancy and postnatal periods” which had a mean ranking of 6.59/10. The mean ranking of each of the top 10 research priorities is shown in Figure 3. The four priority research themes identified were 1) access to healthcare during the COVID-19 pandemic, 2) the direct and 3) indirect effects of COVID-19 on pregnant and breastfeeding women and children and 4) the transmission of COVID-19 and protection from infection.

**Figure 3.**
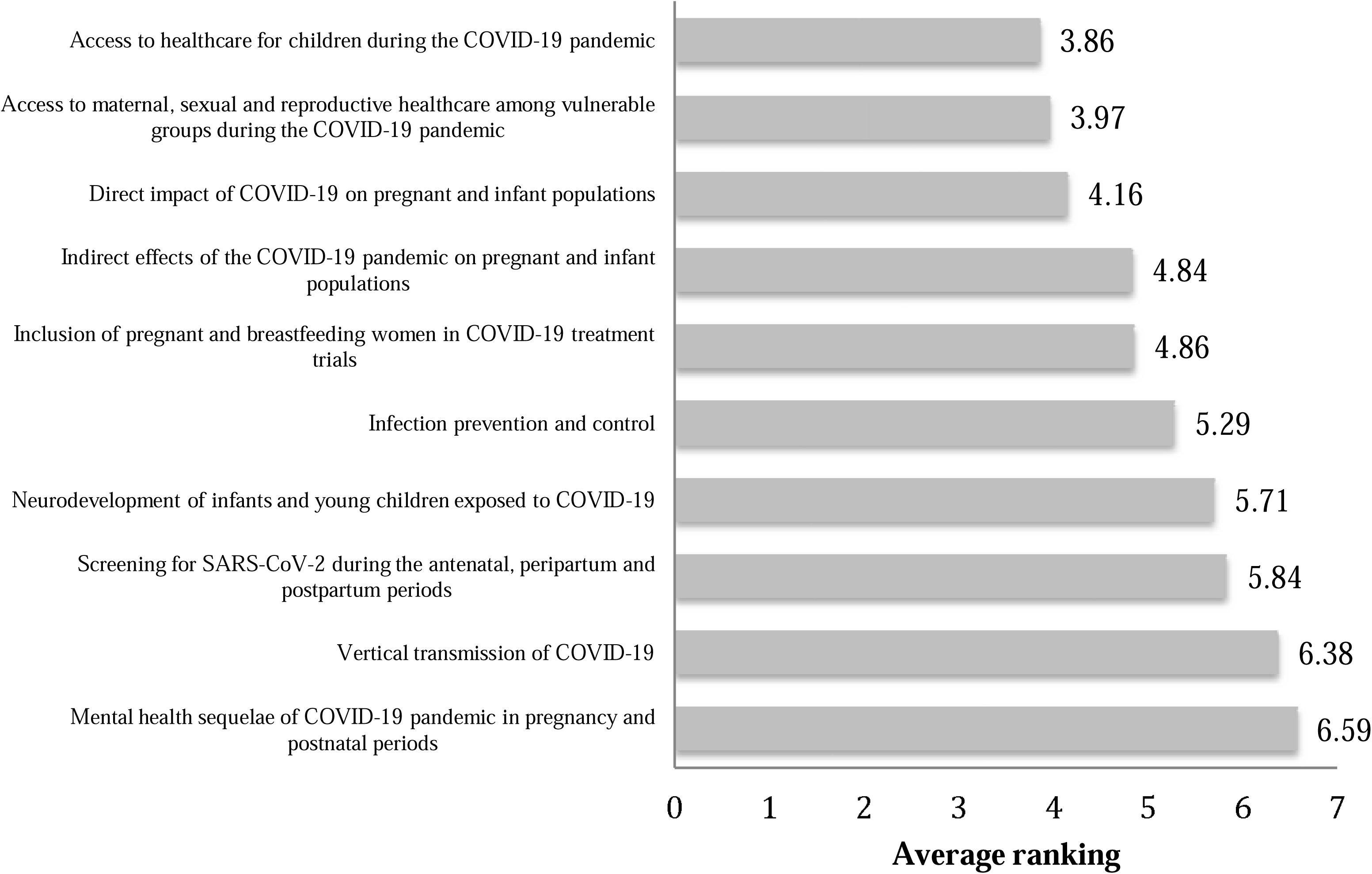
Average rankings of the MNCH research priorities from Questionnaire 2

## Discussion

### Theme 1 Access to healthcare, treatment and vaccine trials for women and children during the COVID-19 pandemic

**Priority research questions**

1. How has the COVID-19 pandemic response affected the availability of and access to maternal, sexual and reproductive health services?
2. What effect has the COVID-19 pandemic response had on the availability of and ability to access child and adolescent health services?
3. What ethical and practical considerations would allow pregnant and breastfeeding women and children equitable access to future COVID-19 treatment and vaccine clinical trials?

The advent of the COVID-19 pandemic has seen unprecedented amounts of pressure being placed on healthcare systems around the world. Acute and critical care services in many countries have been overstretched, leading to the redeployment of many healthcare professionals to the front line and causing a reduction in the provision of “non-urgent” services. In light of this, we must examine what effect these changes have had on women and children their ability to access maternal, sexual and reproductive and child healthcare services during this time. This is particularly pertinent for vulnerable members of society including adolescent girls and unmarried women among migrant and refugee populations who were already facing challenges in accessing these services prior to the onset of the pandemic.(11) Access to healthcare for children during the COVID-19 pandemic was the research priority with the highest average ranking in Questionnaire 2. Evaluating access to maternal and sexual health services for women in vulnerable groups was the most frequently selected research priority among questionnaire respondents based in Africa and was the second most frequently selected research priority among respondents from all other regions, which may reflect concern for a worsening of the pre-existing challenges in the provision of healthcare services that exist due to vast socioeconomic inequalities in many parts of the world.(12)

The inclusion of pregnant and breastfeeding women in COVID-19 vaccine trials was the most frequently selected priority by North American respondents and the fifth most frequently selected research priority among infectious disease specialists who responded to Questionnaire 1. Throughout 2020, pregnant and breastfeeding women were systematically excluded from most COVID-19 treatment and vaccine trials despite urgent calls for their inclusion.(13) Concerns around the lack of safety data also meant that pregnant and breastfeeding women were not prioritized to receive the vaccines after their approval, despite evidence that pregnant women were of greater risk of disease complications.(14,15) In April 2021, the United States Centers for Disease Control and Prevention (US CDC) announced that pregnant and breastfeeding women could safely receive these vaccines after real world data from more than 90,000 pregnant women did not identify any safety signals, however, this came five months after these vaccine were authorized for use in non-pregnant adults.(16–19) While some interventional studies, such as the RECOVERY trial, had widened recruitment to include pregnant and breastfeeding women and children by late 2020, the first large-scale COVID-19 vaccine trial which included pregnant women only commenced recruitment in early 2021.(20) A framework for the ethical and practical considerations of the inclusion of pregnant and breastfeeding women and children in COVID-19 treatment and vaccine trials is urgently needed to ensure that these groups are appropriately considered for inclusion within future treatment and vaccine trials, both in the current pandemic and in future public health emergencies.(21)

### Theme 2 Direct effects of COVID-19 on pregnant and breastfeeding women and children

**Priority research questions**

1. Why are pregnant women with COVID-19 at increased risk of hospitalization, intensive care unit admission and death compared to non-pregnant women? Is there an immunopathological basis for these outcomes?
2. What are the neurodevelopmental effects of exposure to the SARS-CoV-2 virus in utero?
3. What is the immunopathological mechanism of severe COVID-19 in children – pediatric multisystem inflammatory syndrome temporally associated with SARSCoV-2 (PIMS-TS)?

The direct effects of COVID-19 on pregnant women, fetuses and children remain an important area of research as we enter the second year of the pandemic. This research priority was the most frequently selected research priority among OB/GYN and Infectious Diseases respondents and among respondents from Central and South America. Data from the US CDC has shown that pregnant women are at greater risk of intensive care unit admission and death due to COVID-19 than non-pregnant women, and early research suggests that poorer birth outcomes will likely be more prevalent among pregnant women from vulnerable populations within LMICs.(14,15,22–24) Additional observational studies have also identified that women diagnosed with COVID-19 during pregnancy are at higher risk for pre-eclampsia, preterm birth and stillbirth than women who did not have COVID-19 during their pregnancy.(14,25,26) The increased severity of illness among pregnant women observed in the US and UK during the second wave of the pandemic has also raised questions about whether emerging variants of the SARS-CoV-2 virus may be more pathogenic in pregnancy.(27) Greater emphasis should be placed on establishing the pathophysiological and immunological bases for these outcomes and understanding the mechanisms through which these complications occur. Universal screening of pregnant women for COVID-19 may also facilitate greater inclusion of asymptomatic carriers in future studies, which may further improve our understanding of the full breadth of effects of COVID-19 in pregnancy and beyond. Where logistically and financially feasible, efforts should be made to upscale COVID-19 testing to facilitate the identification of women who may be at greater risk of complications.(28)

The impact of COVID-19 on developing fetuses and the manifestation of severe disease in children remain poorly elucidated. Understanding the neurodevelopmental effects of exposure to the SARS-CoV-2 virus on fetuses and infants was deemed to be the highest priority among our pediatric specialist respondents and was the second highest priority among Central and South American respondents.(29,30) Determining the mechanism of the severe form of COVID-19 in children, pediatric multisystem inflammatory syndrome temporally associated with SARS-CoV-2 (PIMS-TS), also known as multisystem inflammatory syndrome in children (MIS-C), was also among the five most frequently selected priorities by pediatric specialist respondents to Questionnaire 1. While observational data from HICs have led to an understanding of potential risk factors for this disorder, including older age and non-white ethnicity (observed in both the UK and South Africa), the immunopathological basis for manifestations of severe disease remains unascertained.(31,32) A large proportion of children diagnosed with PIMS-TS require critical care admission and suffer significant morbidity, including the development of coronary artery aneurysms, shock and multi-organ failure.(31,33) Strengthening our understanding of the causes of this manifestation will help advance therapeutic and management protocols which may help to slow progression of disease.(32)

### Theme 3 Indirect effects of the COVID-19 pandemic on pregnant and breastfeeding women and children

**Priority research questions**

1. What is the effect of the pandemic on rates of sexual and gender-based violence and maternal and child mental health?
2. How will the disruption of routine childhood disease prevention services affect future rates of vaccine preventable diseases?
3. What are the long-term effects of the disruptions in education on childhood development?

Understanding the indirect effects of the COVID-19 pandemic was the most frequently selected research priority among Questionnaire 1 respondents from Europe and among the three most frequently selected research priorities among OB/GYN, sexual and reproductive health, and public health specialists. While COVID-19 has been seen to directly impact the health of pregnant women and their infants, it is also important that we understand the effect that the behavioral and societal responses to the pandemic have had on this cohort. A notable rise in sexual and gender based violence has been documented in many global regions during the pandemic, and while research has identified higher rates of self-reported depression and anxiety among pregnant and postpartum women, a reduction in mental health and support services has also been noted in many parts of the world.(34–37) In light of this, we must seek to better understand the full extent of the challenges these women face in order to design more robust support measures for them as the pandemic persists.

Disruptions in the provisions and access to healthcare services for children, particularly during infancy, may also have unintended consequences on the prevalence of other communicable diseases. Some LMICs have seen disruptions in their ability to provide vaccinations in accordance with their national immunization schedules, owing to both limitations in healthcare resources and the inability of parents and caregivers to bring their children to healthcare facilities due to increased poverty and travel restrictions.(38) This disruption may result in outbreaks of vaccine-preventable diseases in future years, thus, it is important to develop the infrastructures necessary to strengthen immunization programs, facilitate the provision of catch-up vaccinations and where possible, and to monitor disease outbreaks to minimize the re-emergence of such diseases within these communities.(39)

Lockdown measures implemented in many countries have also led to long-term closures of schools and other educational institutions across the world, with remote learning via online platforms replacing face-to-face learning in many countries. The introduction of remote learning has unfortunately served to further disadvantage millions of children from marginalized communities.(40) Long-term longitudinal studies may be necessary to understand how this disruption in schooling has affected child development and objective educational levels in children, particularly among children in resource-limited regions of the world who have faced prolonged restricted access to learning resources during the pandemic.

### Theme 4 Transmission of COVID-19 and protection from infection

**Priority research questions**

1. Is there evidence of mother-to-child transmission of COVID-19? If so, what are the potential routes of transmission?
2. Do maternally-derived antibodies against the SARS-CoV-2 virus confer protective immunity against COVID-19 in infants?
3. What is the evidence for current infection prevention control measures recommended for pregnant and breastfeeding women?

Understanding the risk and routes of transmission of any communicable disease is essential for controlling its spread. Identifying the potential routes of mother-to-child-transmission of COVID-19 is of particular importance for healthcare professionals to be able to provide appropriate care and advice for pregnant and breastfeeding women to minimize the risks posed to their infant. It is well known that some viral infections during pregnancy can have devastating effects on the developing fetus, yet there have been few large-scale studies which involve the collection of samples from pregnant women infected with COVID-19 to determine the routes through which the virus, and immunity to the virus, can be passed from mother to child. There is emerging evidence that suggests that maternally-derived IgG antibodies against the SARS-CoV-2 virus can be transferred to the developing fetus transplacentally.(41) There is also evidence that IgA antibodies against the SARS-CoV-2 virus can also found in the breastmilk, although it is not yet determined whether this is sufficient to confer immunity against COVID-19.(42) These data are urgently needed to better inform infection prevention and control measures and improve care of infants born to mothers infected with the virus during pregnancy and in the postpartum period, a research priority which was among the four most frequently selected research priorities for Asian, African and North American respondents.

The results of the questionnaires indicate a high level of concordance among continents and specialties regarding priority research themes. They highlight the importance of pursuing research which has often been overlooked, including addressing the emotional and psychological impact of the pandemic on maternal and child health, improving access to antenatal, child health services and vaccination during the pandemic, particularly in lower resource settings, as well as investing in long-term follow-up of infants born to women with COVID-19 during the pandemic. Addressing these priorities will require diverse research methodologies, including laboratory-based analyses, qualitative and quantitative research, and population science. The development of generic infrastructure could help foster research collaboration, including the use of core outcome sets, low cost data repositories, and standardized approaches to the reporting of research.(43) We have already seen examples of international collaboration in a number of registries and studies set up to collect data from women who developed COVID-19 during pregnancy, including the COVI-PREG, PAN-COVID and periCOVID studies,(44–46) and in consortiums such as The African coaLition for Epidemic Research, Response and Training (ALERRT) and the COVID-19 Clinical Research Coalition,(47) which aim to facilitate and coordinate COVID-19 research efforts in LMICs. It is hoped the development of a prioritized research agenda could be an important step in further deepening future international research collaboration.

## Limitations

There was an over-representation of Central and South American respondents among the responses to Questionnaire 1. We sought to obtain as many responses from as diverse a group of respondents as possible, however, it is likely that the questionnaire was more successfully disseminated via networks in this region compared to other parts of the world. There was also a preponderance of pediatric infectious diseases societies among the networks through which Questionnaire 1 was disseminated, which may have skewed responses received towards pediatric infectious diseases specialists. We chose to disseminate the questionnaires through affiliated memberships of the working group members and via social media as we believed this would allow a greater inclusion of clinicians and frontline workers as well as researchers in resource-limited nations. In disseminating the questionnaires using these links, it is possible that our results may have been subject to sampling bias. To mitigate the effect of these potential biases, we performed a subgroup analysis to ensure that the views of respondents from all regions and specialties were equitably reflected among the final list of research priority themes.

There was also a significantly reduced response rate to Questionnaire 2 (21.7% of the original pool of respondents), possibly as the dissemination of our second questionnaire coincided with the second wave of the COVID-19 pandemic in many of the countries where our respondents were based.(48–50) This reduction in engagement may have therefore reflected local challenges faced by the clinicians and researchers due to an increasing number of COVID-19 cases, although we cannot exclude other causes for this reduced response rate. Given the degree of attrition, we considered the subgroup preferences from Questionnaire 1 when developing the final research priority themes to ensure parity among responses and to lessen the influence of the reduction in response rate on the final research priorities derived from the questionnaire data. In light of the low response rate to Questionnaire 2, it was also decided that a modified Delphi approach would be used with only two questionnaire rounds. Given that there was a high degree of consensus among the answers to Questionnaire 1, we believe this method remained appropriate for the development of these four research priority themes.

Although space was provided for additional comments from respondents, the use of questionnaires imposed limitations on the depth of the data we were able to collect. While the questionnaires were useful for collecting data from a large number of professionals within our target field, we were limited in our ability to further explore the response we received. Future work may require a more in-depth exploration of ideas through interviews and focus group discussions with respondents to further expand the responses given in the questionnaires to better understand the perceived research priorities within different global regions.

## Conclusion

A prioritized list of research uncertainties, developed to specifically highlight the most pressing clinical and research needs as perceived by healthcare professionals and researchers in both HICs and LMICs should help funding organizations and researchers to answer pertinent questions related to pregnancy and childhood during the pandemic. Although there was a great degree of consensus among the priorities selected by our questionnaire respondents, it is important that an approach is undertaken which also sought to identify differences in international research needs. The selected list of research uncertainties should serve to focus discussion regarding the allocation of limited resources and priorities should be reviewed on a regular basis to reflect the evolving availability of evidence.

While global research collaboration is of great importance in furthering our collective understanding of the effects of COVID-19 on pregnant and breastfeeding women and children, it is essential that this research also serves the needs of the population in which the research is conducted and provides data relevant to their needs. We hope that the findings of this study will support researchers and policy makers worldwide to better understand the needs of their region, enabling the prioritization of research that aligns with the priorities of their communities.

## Data Availability

Data is available upon request.

## Authorship statement

ME^1^, JA, RS, TR, ME^2^, KGD, YA and KLD drafted the questionnaires. ME^1^ and KLD analyzed the questionnaire data. MNE drafted the first version of the manuscript. ME^1^, JA, SPS, RS, TR, ME^2^, KGD, TA, YA, AC, DE, AK, KLD contributed equally towards the revising and editing of the manuscript. All authors approved the final version of the manuscript prior to submission.

## Appendices

## Appendix 1

### Questionnaire 1

The Maternal, Newborn and Child Health (MNCH) Working Group of the COVID-19 Clinical Research Coalition would like to seek your opinion about priority research areas relating to COVID-19 and MNCH. We would be grateful for your answers to the questions in this short survey. Thank you.

1. Your specialty:
  - Obstetrics and Gynecology
  - Pediatrics
  - Sexual and Reproductive Health
  - Midwifery and Nursing
  - Infectious Diseases/Pathological Sciences
  - Public Health
  - Epidemiology
  - Other
2. Institution (including country):
3. Are you currently involved in research related to COVID-19? Y/N
  a. If yes, what is your main area of focus?
4. Please select the five most urgent COVID-19 MNCH research areas from the list below
  - Access to maternal, sexual and reproductive healthcare among vulnerable groups during the COVID-19 pandemic
  - Access to healthcare for children during the COVID-19 pandemic
  - Direct impact of COVID-19 on pregnant and infant populations
  - Immunological mechanisms of resilience against COVID-19 among children
  - Inclusion of pregnant women in COVID-19 vaccine trials
  - Inclusion of pregnant and breastfeeding women in COVID-19 treatment trials
  - Indirect effects of the COVID-19 pandemic on pregnant and infant populations
  - Infection prevention and control
  - Mental health sequelae of COVID-19 pandemic in pregnancy and postnatal periods
  - Neurodevelopment of infants and young children exposed to COVID-19
  - Pathophysiology and long term sequelae of pediatric inflammation and multisystem syndrome (PIMS-TS)
  - Pathophysiological mechanisms of severe disease in children
  - Potential aerosolization of SARS-CoV-2 during second stage of labor
  - Prevention and treatment strategies for pregnant women and newborns in humanitarian settings
  - Safe breastfeeding
  - Screening for SARS-CoV-2 during the antenatal, peripartum and postpartum periods
  - Vertical transmission of COVID-19

6. Please provide any other MNCH priority research areas which were not included in the list
7. Any additional comments?
8. Would you be happy to be contacted by a representative of the Maternal, Newborn and Child Health Working Group about your answers to this survey? Y/N

If yes, please provide

Name

Email address

## Appendix 2

### Questionnaire 2

Thank you for agreeing to be contacted about the results of the MNCH Research Priorities questionnaire sent by the COVID-19 Clinical Research Coalition earlier this year.

We have reviewed the responses of the first questionnaire and identified the highest ranked research priorities. We would be grateful for additional feedback to refine these priorities further. Thank you.

1. Please rank the 10 research priorities from the list below (MOST important = 1, LEAST important=10)
  1. Access to maternal, sexual and reproductive healthcare among vulnerable groups
  2. Direct impact of COVID-19 on pregnant and infant populations
  3. Indirect effects of the COVID-19 pandemic on pregnant and infant populations
  4. Mental health sequelae of COVID-19 pandemic in pregnancy and postnatal periods
  5. Access to healthcare for children during the COVID-19 pandemic
  6. Infection prevention and control
  7. Inclusion of pregnant and breastfeeding women in COVID-19 treatment trials
  8. Vertical transmission of COVID-19
  9. Prevention and treatment strategies for pregnant women and newborns in humanitarian settings
  10. Screening for SARS-CoV-2 during the antenatal, peripartum and postpartum periods
2. Do you think these 10 priorities accurately reflect the MNCH research priorities in your region? Y/N
  - If no, please provide details of additional research priorities which should be considered:
3. Are the research priorities understandable in their current format? Y/N
  - If no, please provide comment:
4. Any additional comments? Y/N
5. We plan to publish the results of this questionnaire in a scientific journal. Would you like to be named within the author group of this paper? Y/N
  - If yes, please provide the following details:

Title

Full name

Institutional affiliation:

## Appendix 3

### Location (countries) of Questionnaire 1 respondents

Afghanistan

Argentina

Belgium

Benin

Burkina Faso

Colombia

Costa Rica

Cuba

Democratic Republic of Congo

Honduras

India

Iraq

Japan

Madagascar

Malawi

Mali

Mexico

Niger

Pakistan

Peru

Saudi Arabia

South Africa

Sweden

Switzerland

Tunisia

United Kingdom

Unites States of America

Uganda

Venezuela

